# A scoping review identified additional considerations for defining estimands in cluster randomised trials

**DOI:** 10.1101/2025.06.25.25330127

**Authors:** Dongquan Bi, Andrew Copas, Fan Li, Brennan C Kahan

## Abstract

**Background and objective:** An estimand is a clear description of the treatment effect a study aims to quantify. The ICH E9(R1) addendum lists five attributes that should be described when defining an estimand. However, the addendum was primarily developed for individually randomised trials. Cluster randomised trials (CRTs), in which groups of individuals are randomised, have additional considerations for defining estimands, such as how individuals and clusters are weighted. We aimed to identify a list of additional items that may need to be considered when defining estimands in CRTs.

**Study design:** We conducted a systematic search of multiple databases as well as the authors’ personal libraries to identify articles that described an aspect of an estimand definition for CRTs which was not explicitly covered by one of the five attributes listed in the ICH E9 (R1) addendum. From this, we generated a list of items that may require consideration when defining estimands for CRTs beyond the five attributes listed in the ICH E9(R1) addendum.

**Results:** From 46 eligible articles, we identified 8 items that may need to be considered when defining estimands in CRTs: (i) population of clusters; (ii) population of individuals under selection bias; (iii) exposure time of individuals/clusters on treatment; (iv) how individuals and clusters are weighted (e.g. individual-average vs. cluster-average); (v) whether summary measures are marginal or cluster-specific; (vi) strategies used to handle cluster-level intercurrent events; (vii) how interference/spillover is handled; and (viii) how individuals who leave or change clusters are handled.

**Conclusion:** This review has identified additional items that may need to be considered when defining estimands for CRTs. Study investigators undertaking CRTs should consider these items when defining estimands for their trials, to ensure estimands are unambiguous and relevant for end-users such as clinicians, patients, and policy makers.

**What is new?:** *Key findings:*

- The review identified eight additional items that are not explicitly described by the existing attributes listed in the ICH E9(R1) addendum, but that may need to be considered when defining estimands for cluster randomised trials

*What this adds to what is known:*

- Defining estimands using the attributes set out in the ICH E9(R1) addendum can promote clarity, however these attributes alone are not sufficient to describe a clearly defined research question in cluster randomised trials
- In addition to the five attributes set out in ICH E9(R1), investigators conducting cluster randomised trials need to consider which additional items are relevant to describe when defining their estimand

*What is the implication and what should we change now:*

- Items identified in this review can be used by investigators conducting cluster randomised trials to help define their estimand to ensure it is clear
- Specific guidance on defining estimands for cluster randomised trials is required to ensure that research questions are fully described in these trials

## 1. Introduction

An estimand is a clear description of the treatment effect a trial aims to quantify [1, 2]. The use of estimands can help to clarify the question being investigated and ensure statistical methods are well-aligned to the research question. In 2019, the International Council for Harmonisation published the ICH E9(R1) addendum [1], which sets out a framework for defining an estimand. This involves specifying five attributes: (1) population of patients; (2) treatment conditions being compared; (3) endpoint/outcome variable; (4) population-level summary measure; and (5) strategies used to handle intercurrent events (post-randomisation events that affect interpretation or existence of the endpoint). [1]

The ICH E9 (R1) addendum was developed by regulators and the pharmaceutical industry, primarily with individually randomised trials in mind. Although the five attributes defined within the addendum will be relevant for all study designs, some designs may require specification of additional considerations in order to have a well-defined estimand. For example, cluster randomised trials (CRTs) involve randomising groups of individuals to different interventions. Because the type of cluster may affect how well the intervention works (e.g. a free-school lunch programme may provide more benefit in public schools than in private schools), specification of the population of clusters, in addition to the population of individuals, may be required for the estimand definition.[3-8] Other issues have been noted in the literature, including specification of how individuals and clusters are weighted (e.g. whether interest lies in the individual-average or cluster-average treatment effect) [5, 8, 9] and how potential outcomes are summarised and contrasted (e.g., whether interest lies in the marginal or cluster-specific effect) [5, 8].

Failure to address these additional items in CRTs could lead to ambiguous estimands, which in turn may cause misalignment of statistical estimation methods with the intended estimands and misinterpretation of results. There is a need to provide guidance on any additional items that need to be considered when defining estimands for CRTs that are not explicitly listed in the ICH E9 (R1) addendum. We therefore conducted a scoping review to identify a list of additional items that may need to be considered when defining estimands in CRTs.

## 2. Methods

We conducted a scoping review to identify articles that described unique considerations for estimands in CRTs. The aim was to produce a list of additional items that may require consideration when defining estimands in these trials.

### 2.1. Eligibility Criteria

Articles were eligible if they described an aspect of an estimand definition for CRTs which was not explicitly covered by one of the five attributes listed in the ICH E9 (R1) addendum.

### 2.2. Search strategy

We used the following sources to identify eligible papers: (1) a systematic search of EMBASE, MEDLINE, Scopus, and the Web of Science databases, and (2) relevant papers from the authors’ personal libraries.

For the database searches, the search strategy was developed in consultation with a research librarian. Two searchable concepts “ estimands” and “ cluster randomised trials” were chosen. For each concept, we identified synonyms and alternative terminology. The search required results to contain terms for both concepts. The full search strategy can be found in Appendix 1 in the Supplemental File.

The search was performed in June 2024. One author (DB) assessed potential eligibility from the title and abstracts of identified articles and undertook full-text review to confirm eligibility. Queries regarding eligibility were discussed with at least one other author (BCK and/or AC).

### 2.3. Data extraction

A data exaction form was developed and piloted (see Appendix 2 in the Supplemental File). In addition to a brief description of the article, we extracted information on any additional items that may need to be described when defining estimands for CRTs that were discussed in the article.

One author completed the extractions (DB). Queries were discussed with at least one other author (BCK and/or AC), with disagreements resolved by discussion.

### 2.4. Analysis

Data analysis was primarily descriptive. Any considerations that were relevant to the estimand definition for CRTs were recorded. This generated a list of potential items that may require consideration when defining estimands for CRTs beyond the five attributes listed in the ICH E9(R1) addendum. Results were reported in line with the PRISMA extension for scoping reviews.[10]

## 3. Results

### 3.1. Search results

The database search identified 1294 articles (Figure 1). After duplicates were removed, 613 articles underwent title and abstract screening, which resulted in 582 articles being excluded. The full text of the remaining 32 articles was screened for eligibility. After full-text screening, 24 articles were found to be eligible.

**Figure 1.**
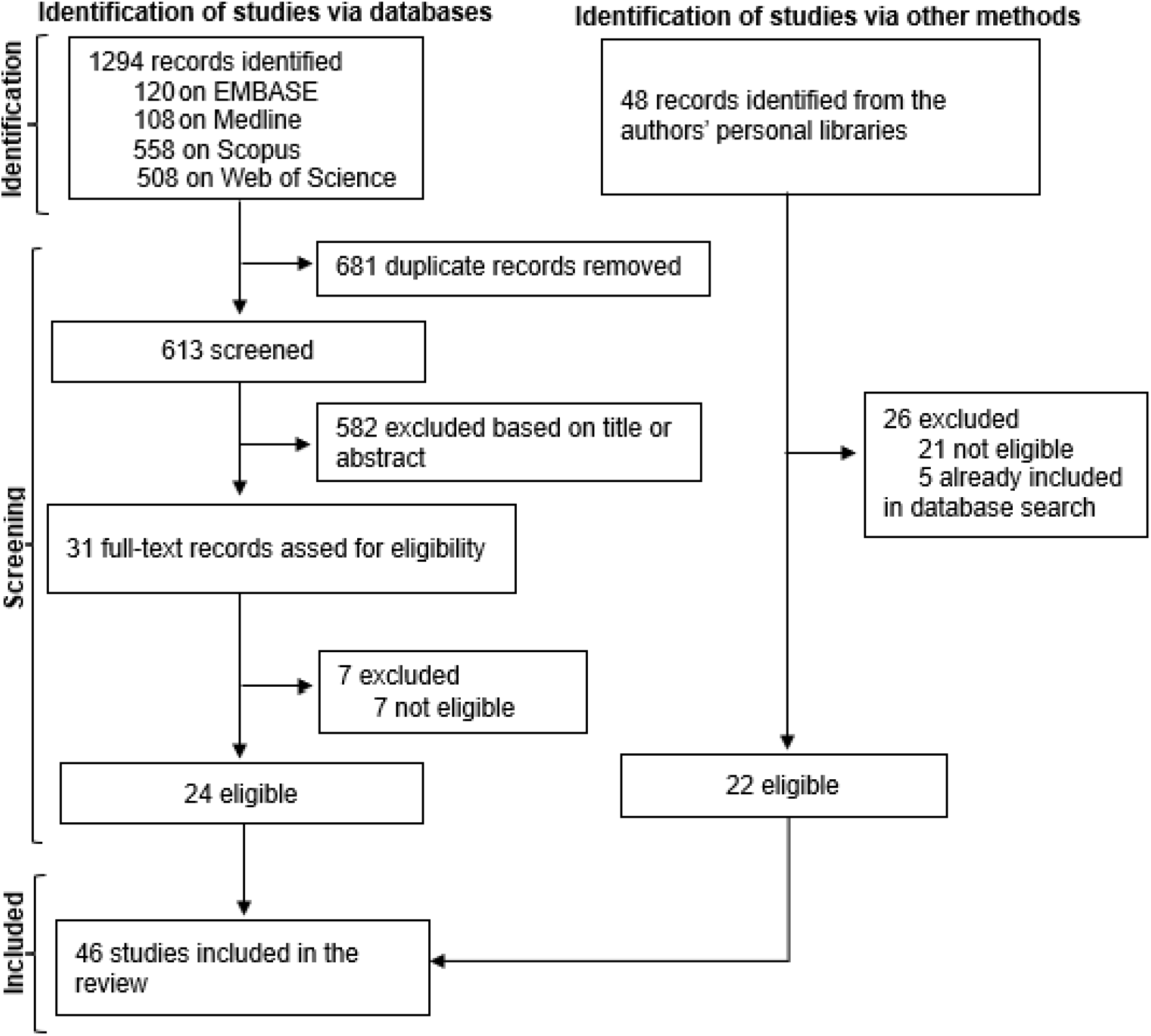
Flow diagram of the search process

An additional 48 articles were identified from authors’ personal libraries. After full-text screening, 26 articles were excluded either because they were duplicates of articles identified in the database search, or did not meet the eligibility criteria. This left 22 eligible articles.

Combining the two sources resulted in 46 articles that were eligible to be included in this review (Figure 1). The full list of these articles can be found in Appendix 3 in the Supplemental File.

### 3.2. Additional items that may need to be described for estimands in CRTs

The review identified 8 additional items that may need to be considered when defining estimands in CRTs (Table 1):

**Table 1.**
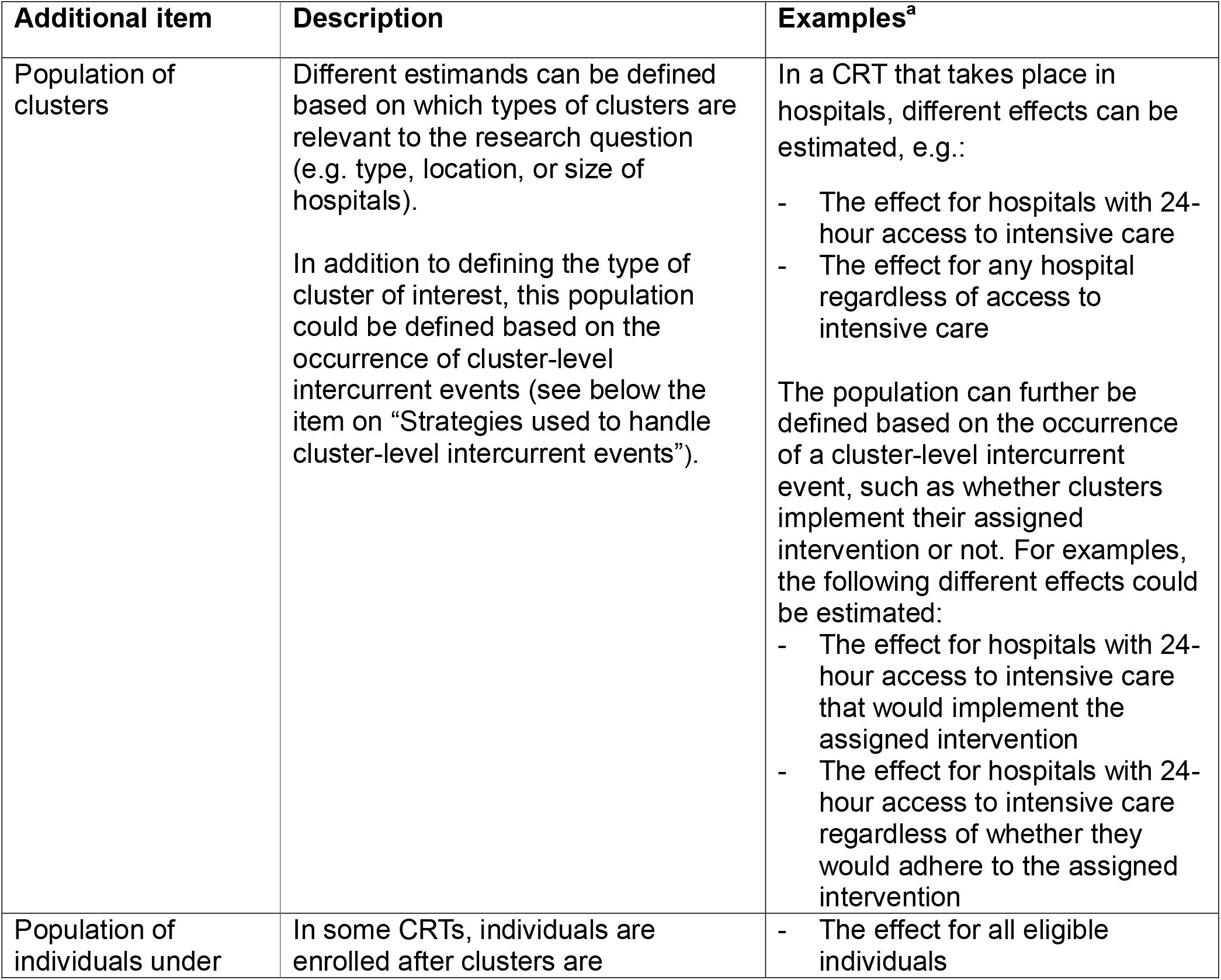
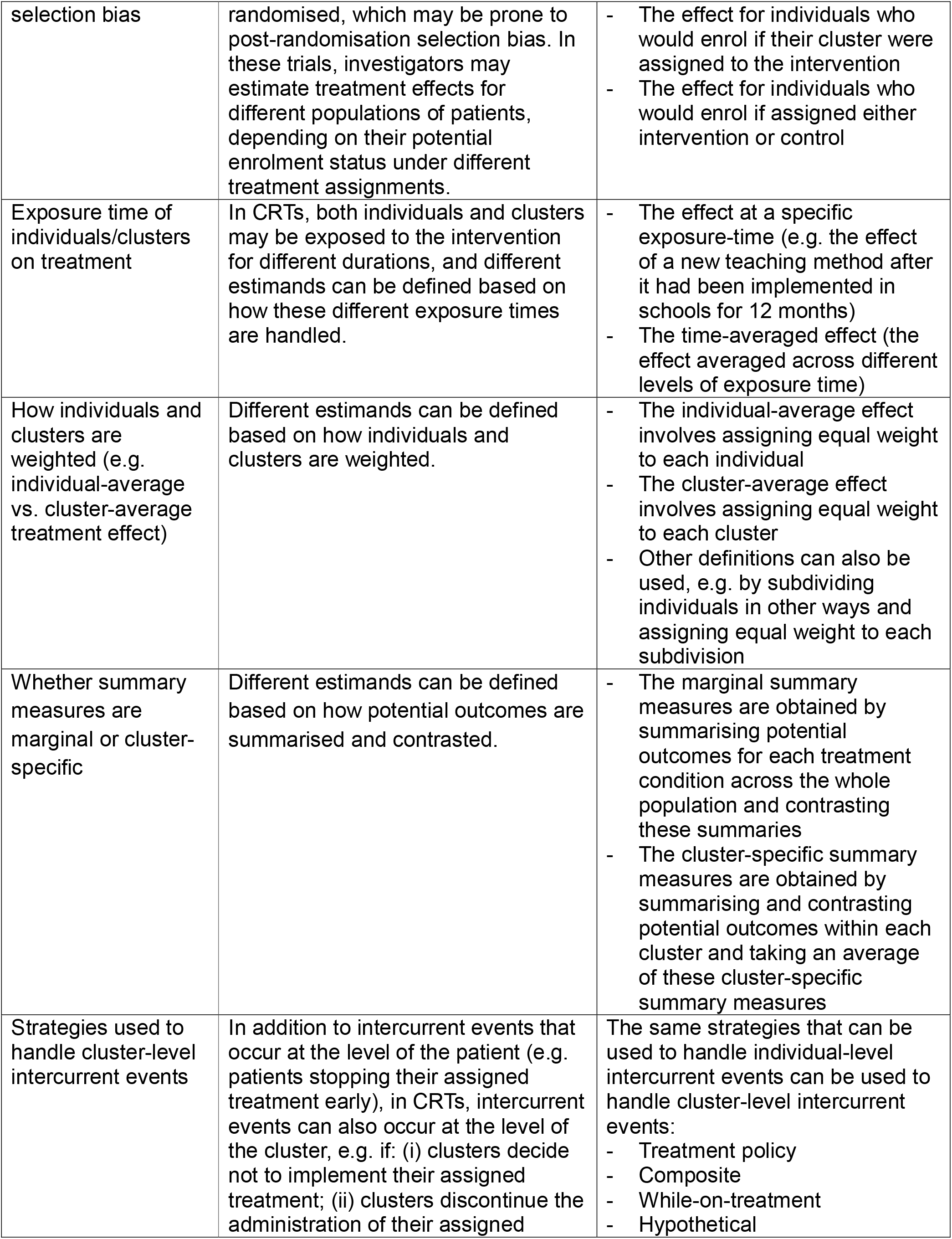
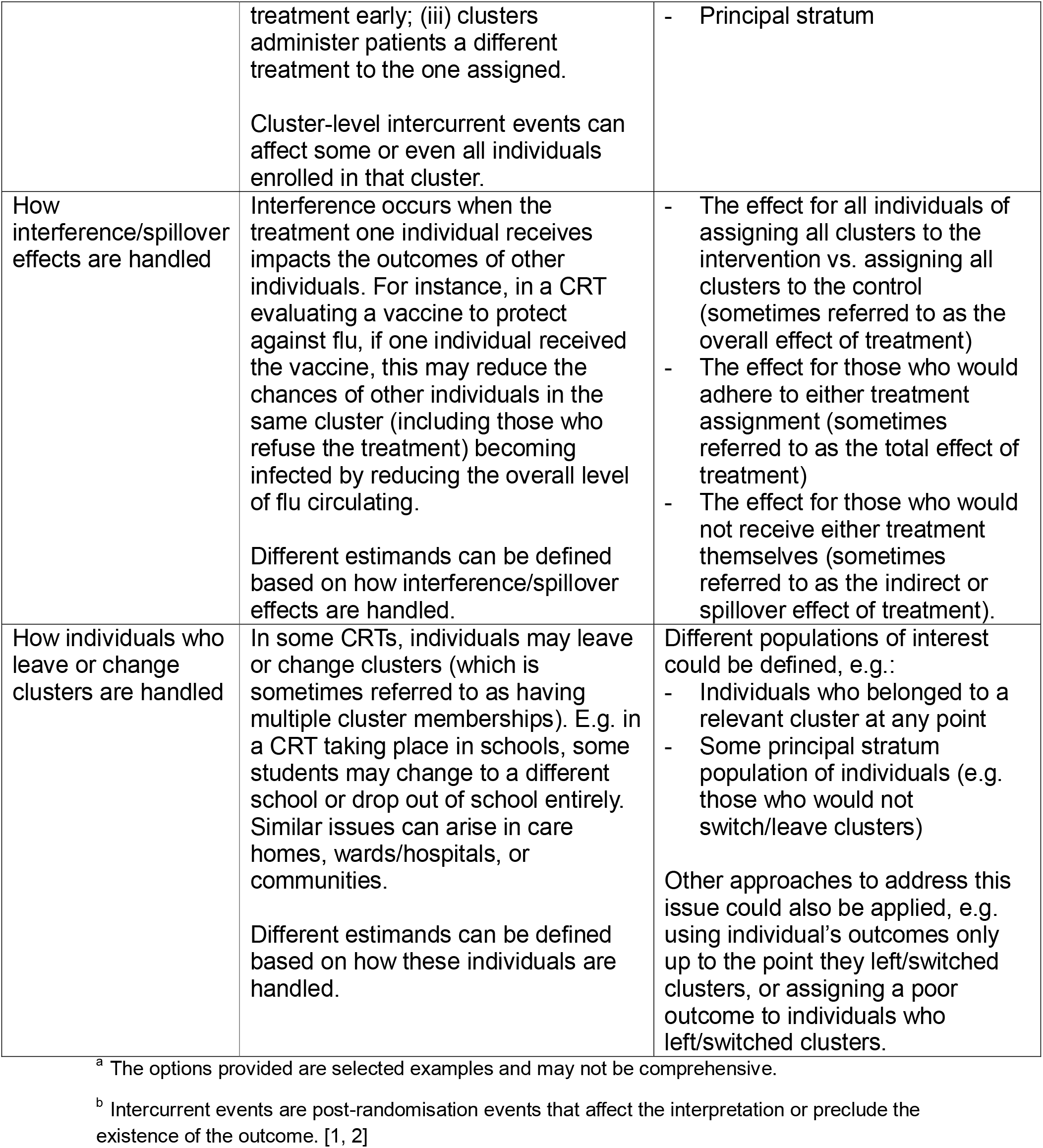
List of additional items which may require consideration when defining estimands for cluster randomised trials.

- Population of clusters [5, 7, 11]
- Population of individuals under selection bias[3, 4, 6, 12, 13]
- Exposure time of individuals/clusters on treatment [14-18]
- How individuals and clusters are weighted (e.g. individual-average vs. cluster-average) [7, 12, 19-32]
- Whether summary measures are marginal or cluster-specific [5, 11, 27, 28, 33-39]
- Strategies used to handle cluster-level intercurrent events [5, 40-43]
- How interference/spillover is handled [24, 25, 39, 44-50]
- How individuals who leave or change clusters are handled [51]

## 4. Discussion

In this scoping review, we set out to identify a list of additional items that may need to be considered when defining estimands for CRTs. After reviewing 46 articles, we found 8 additional items that may be relevant for CRTs. These items highlighted that for many CRTs there may be a need to further clarify certain aspects of the estimand, including aspects related to the population, treatment conditions, summary measure, and strategies to handle intercurrent events.

For example, this review identified the need to further clarify the populations of interest in CRTs, including which individuals are part of the population of interest when there is selection bias, and which population of clusters is of interest; how intercurrent events at the level of the cluster, and how individuals who leave or change clusters, will be handled; how summary measures will be defined (e.g. whether individual-vs. cluster-average and marginal vs. cluster-specific effects are of interest); and how exposure time on treatment and interference/spill-over effects will be handled, when applicable.

The items identified in this review can be used by investigators conducting CRTs to evaluate whether additional items should be considered when defining the estimand for their study, over and above the existing attributes listed in the ICH E9(R1) addendum. Consideration of these additional items could lead to less ambiguous estimands which make study results easier to interpret for end-users, such as clinicians, policy makers, and patients, and provide a clearer guide for investigators themselves on how to design and analyse their trial in line with study objectives. However, it should be noted that not all items will be relevant for all studies. For instance, some items, such as how estimands are defined when there is selection bias, or interference/spillover effects, will only be relevant in situations when these specific issues arise.

This study highlights some important areas of future research. The ICH E9(R1) addendum provides important guidance for defining estimands, however, additional guidance for defining estimands in CRTs is essential to ensure estimands are clear and unambiguous in these trials. A consensus-extension of the ICH E9(R1) addendum for CRTs is currently underway [52], and the results from this scoping review will play an essential role in highlighting which additional items or attributes could be considered for inclusion in such guidance. Secondly, although there has been a wide range of methodological work around methods to estimate different estimands, it would be useful to collate such information together in tutorials or overview papers, to provide investigators with a clear list of options around which analyses could be implemented for specific estimands, along with the required assumptions of each analysis method.

Strengths of this study include a comprehensive search of multiple databases and inclusion of articles from the authors’ personal libraries. However, we acknowledge that single reviewer screening could have resulted in some eligible articles being missed and that single data extracting could have resulted in information not being extracted comprehensively. To mitigate this risk, however, the final extraction results were checked by another reviewer (BK) which should help to minimise errors. Furthermore, due to the scoping nature of the search and the purpose of the review being to identify concepts that are relevant to the estimand definition for CRTs, single data extraction was deemed sufficient for this objective.

## 5. Conclusion

This review has identified additional items that may need to be considered when defining estimands for CRTs. Study investigators undertaking CRTs should consider these items when defining estimands for their trials, to ensure estimands are unambiguous and relevant for end-users such as clinicians, patients, and policy makers.

## Data Availability

All data produced in the present work are contained in the manuscript

## CRediT authorship contribution statement

Dongquan Bi: Writing – original draft, Writing – review & editing, Data curation, Formal analysis, Methodology.

Brennan Kahan: Writing – review & editing, Conceptualization, Supervision, Methodology.

Andrew Copas: Writing – review & editing, Conceptualization, Supervision, Methodology.

Fan Li: Writing – review & editing, Methodology.

## Acknowledgments

We thank Dr. Debora Marlett, the Training and Clinical Support Librarian from UCL Library Services for her advice on developing the search strategy.

